# Precision Stratification: Kinase Assay in Asian *LRRK2* Risk Carriers and Idiopathic Parkinson Disease

**DOI:** 10.1101/2025.09.23.25336067

**Authors:** Tzi Shin Toh, Lei Cheng Lit, Shen-Yang Lim, Jia Wei Hor, Choey Yee Lew, Anis Nadhirah Khairul Anuar, Yi Wen Tay, Kirsten Black, Jia Lun Lim, Jannah Zulkefli, Kai Shi Lim, Hans Xing Ding, Shalini Padmanabhan, Azlina Ahmad-Annuar, Eng King Tan, Dario R. Alessi, Esther Sammler, Ai Huey Tan

## Abstract

**Importance:** LRRK2 kinase inhibition is one of the most promising therapeutic strategies in Parkinson’s disease (PD), yet understanding of kinase activity in the Asian-prevalent p.G2385R and p.R1628P variants, each affecting 5–10% of PD patients, remains limited. Development of patient stratification and target engagement markers applicable across global cohorts is an urgent priority.

**Objective:** To investigate pRab10^Thr73^ phosphorylation and pLRRK2^Ser935^ dephosphorylation as markers of *in vivo* LRRK2 activation status in human monocytes from manifesting carriers of p.G2385R and p.R1628P, and idiopathic PD (iPD), with clinical correlations.

**Design:** Cross-sectional, observational study conducted between 2021 to 2024.

**Setting:** Multi-ethnic Asian cohort from two quaternary hospitals in Malaysia.

**Participants:** This study included 242 participants, consisting of PD-G2385R (*n*=57), PD-R1628P (*n*=61), PD-G2385R+R1628P (*n*=5), as well as age- and sex-matched iPD (*n*=61) and healthy controls (HC; *n*=58) who were negative for p.G2385R and p.R1628P. Exclusion criteria included recent acute infections and inability to complete assessments.

**Exposure:** PD with and without *LRRK2* p.G2385R/p.R1628P risk variants.

**Main Outcomes and Measures:** pRab10^Thr73^ and pLRRK2^Ser935^ phosphorylation were measured using multiplexed quantitative immunoblotting. Clinical severity was evaluated using standardized MDS-UPDRS, CISI-PD, and MoCA ratings.

**Results:** Compared to HC, pRab10^Thr73^ phosphorylation was significantly elevated in PD-G2385R (∼1.2-fold, *p*=0.011) and markedly higher in double-variant carriers (∼2.8-fold, *p*=0.008), but not in PD-R1628P or iPD. In parallel, pLRRK2^Ser935^ phosphorylation was reduced in all PD subgroups (lowest in double-variant carriers) vs. HC, and inversely correlated with pRab10^Thr73^ (*r*_*s*_=-0.611, *p*<0.001). All double-variant carriers, the majority of single-variant carriers, and one-third of iPD exceeded the HC median in pRab10^Thr73^ phosphorylation. Higher pRab10^Thr73^ levels correlated with better cognition, but motor or disability associations were not significant after covariate adjustment.

**Conclusions and Relevance:** LRRK2 kinase activity is enhanced in PD patients with *LRRK2* p.G2385R, with more pronounced effects in those who carry concomitant p.R1628P variant. Elevated kinase activity in a considerable subset of iPD underscores the broader relevance of LRRK2 signaling and therapeutics beyond coding variants. The observed inter-individual variability highlights the role of additional genetic and environmental modifiers and supports biochemical stratification beyond genotyping in future LRRK2 trials.

## INTRODUCTION

The lack of reliable biomarkers for early detection, progression monitoring, and treatment stratification remains a major barrier to precision medicine in Parkinson’s disease (PD).^1^ Recent genetic discoveries are beginning to close this gap by uncovering molecular pathways that can be targeted therapeutically.^2,3^ Among these, the leucine-rich repeat kinase 2 (*LRRK2*) gene stands out as one of the commonest and clinically significant genetic factors in PD.^4^ Importantly, LRRK2 dysfunction has also been implicated in sporadic/idiopathic PD (iPD) (beyond familial cases), underscoring its central role in disease biology.^2,4^ As aberrant kinase function is a hallmark of pathogenic *LRRK2* variants, LRRK2 kinase activity has become a focal point for translational research, with kinase inhibition now considered one of the most promising therapeutic strategies entering clinical trials.^5,6^ An urgent priority is therefore the development of robust LRRK2 kinase assays that can enhance patient stratification (including in iPD) and serve as target engagement markers in LRRK2 therapeutic trials.

Early approaches to measuring LRRK2 activity in human biosamples relied on phosphorylation at autophosphorylation sites (e.g., Ser1292^7,8^), which were limited by low intrinsic levels, or at heterologous sites (e.g., Ser935^9-11^). Recent work demonstrated that phosphorylation of LRRK2 at Ser935 (pLRRK2^Ser935^) reflects its structural/conformational state. When LRRK2 is inactive, Ser935 is highly phosphorylated, allowing interaction with 14-3-3 stabilizing proteins. Upon activation, through pathogenic mutations or type 1 kinase inhibitors that enforce an active conformation, 14-3-3 biding is disrupted, resulting in Ser935 dephosphorylation. Thus, reduced pLRRK2^Ser935^ phosphorylation indicates a shift toward an active LRRK2 conformation.^12^ Consistent with this, reduced pLRRK2^Ser935^ phosphorylation has been observed in various kinase-activating mutations.^13,14^ A breakthrough came with the identification of Rab GTPases as direct LRRK2 substrates.^15^ Among these, Rab10 phosphorylation at Thr73 (pRab10^Thr73^) has emerged as a robust readout of kinase hyperactivation and a pharmacodynamic marker of kinase inhibition across preclinical models and *ex vivo* patient studies.^5,13,15,16^ Despite high Rab10 expression in neutrophils and monocytes and its strong potential as a clinically actionable assay for LRRK2-targeted trials, studies in human biosamples remain extremely limited. To date, pRab10^Thr73^ phosphorylation has only been examined in a small number of *LRRK2* variant carriers, including p.G2019S (*n*=21) and p.R1441G (*n*=21),^17^ as well as for functional validation of rare pathogenic variants including p.F1700L (*n*=1),^18^ p.R1067Q (*n*=1),^19^ and p.V1447L (*n*=1).^20^

Much of our current understanding of LRRK2 biology derives from European cohorts, particularly those with the p.G2019S variant.^2^ In contrast, data on Asian-prevalent *LRRK2* variants, such as p.G2385R and p.R1628P, remain scarce. With each having prevalence rates of 5–10% in Asian PD populations, these variants represent the most important genetic risk factors in East and Southeast Asian individuals (Chinese, Korean, Japanese, Malay, Thai, and Vietnamese).^2,21,22^ Yet their effects on LRRK2 kinase activity remain poorly understood and it is unclear whether they drive the same pathophysiological mechanisms as p.G2019S and other canonical mutations. *In vitro* and *in vivo* models have yielded conflicting results, ranging from reduced to elevated kinase activity.^13,23-29^ Critically, human data are almost entirely absent. This gap is particularly concerning given the projected exponential rise in PD prevalence across Asia in the coming decades.^30^ Without validated assays in these populations, Asian PD patients risk being underserved in ongoing and future LRRK2-targeted clinical trials. Moreover, understanding the kinase biology of these variants could illuminate broader mechanisms by which LRRK2 contributes to iPD, since p.G2385R and p.R1628P are thought to modulate kinase activity in ways that may extend beyond what occurs in monogenic disease.

In this study, we sought to bridge this critical gap by employing a multiplexed immunoblotting assay to measure LRRK2-dependent pRab10^Thr73^ and pLRRK2^Ser935^ phosphorylation in human peripheral blood monocytes, isolated from PD patients carrying the p.G2385R and/or p.R1628P variants, alongside iPD and healthy controls (HC). Additionally, we examined the correlations of these markers with key clinical variables, across the PD cohort and within each PD subgroup.

## METHODS

### Participant recruitment and clinical assessment

The study was approved by the Universiti Malaya Medical Center (UMMC) Medical Research Ethics Committee (MECID No. 2022427-11195), and written informed consent was obtained from all participants. All procedures were conducted in accordance with the Declaration of Helsinki. Manifesting *LRRK2* p.G2385R (PD-G2385R), p.R1628P (PD-R1628P), and double-variant carriers (PD-G2385R+R1628P), as well as iPD, were recruited from the UMMC and Universiti Malaya Specialist Center (UMSC) Neurology Clinics. All patients were diagnosed with PD by movement disorder specialists (SYL and AHT) according to standard clinical criteria.^31^ Spouses of PD patients who tested negative for *LRRK2* p.G2385R and p.R1628P were recruited as HCs. Other inclusion criteria were age >18 years old and available *LRRK2* p.G2385R and p.R1628P genotype status confirmed by TaqMan allelic discrimination assays, as previously described.^21^ Exclusion criteria were inability to complete study assessments and active/recent acute infections.

Clinico-demographic data were collected, including the International Parkinson and Movement Disorder Society-Unified PD Rating Scale (MDS-UPDRS), Hoehn and Yahr staging, and Clinical Impression of Severity Index for PD (CISI-PD), by two movement disorder specialists (SYL and AHT).^21,32^ Cognitive function was evaluated using Montreal Cognitive Assessment (MoCA) by trained research personnel.

### Human monocyte isolation and multiplexed quantitative immunoblotting

Twenty mL fresh blood was collected from participants using EDTA blood tubes. Within an hour of sample collection, monocytes were isolated from the blood samples using the EasySep™ Direct Human Monocyte Isolation Kit (STEMCELL Technologies, Vancouver, Canada, Cat#19669), according to a standardized protocol.^19^ Monocyte lysates were snap-frozen and stored at -80°C until further processing. Multiplexed quantitative immunoblotting was performed for pRab10^Thr73^, pLRRK2^Ser935^, total Rab10, total LRRK2, and GAPDH, as previously described.^13,17^ Detailed methods for monocyte isolation and immunoblotting are provided in Supplementary Methods. The list of primary and secondary antibodies used in this assay is provided in Supplementary Table 1, and representative immunoblots are shown in Supplementary Figure 1.

### Screening for other potential LRRK2-activating variants

All participants were screened for known pathogenic or likely pathogenic variants in *LRRK2* (p.R1067Q, p.N1437H, p.R1441C/G/H, p.Y1699C, p.G2019S, and p.I2020T), *VPS35* (p.D620N), and *RAB32* (p.S71R),^13,33^ using the NeuroBooster Array (NBA) conducted through the Global Parkinson’s Genetics Program.^34^ Genotyping data were extracted and analyzed via PLINK 2.0.^35,36^

### Statistical analysis

Statistical analyses were performed using R version 4.4.0. The Shapiro-Wilk test was used to determine if the data followed a normal distribution. Between-group differences were evaluated using analysis of variance (ANOVA) for normally distributed data, the Kruskal-Wallis test for non-normally distributed data, and the chi-square test for categorical data. Pairwise comparisons of the ratios of pRab10^Thr73^/total Rab10, total Rab10/GAPDH, pLRRK2^Ser935^/total LRRK2, and total LRRK2/GAPDH were conducted using the Kruskal-Wallis test with post hoc Dunn’s test, with *p*-values adjusted using the Benjamini-Hochberg method. Spearman’s correlation analysis was used to evaluate the relationships between pRab10^Thr73^/total Rab10 or pLRRK2^Ser935^/total LRRK2 and various clinical variables, both in the overall PD group and within each PD subgroup. Generalized linear models were used to control for potential covariates (i.e., age, age at diagnosis, or disease duration) in analyzing these clinical associations.

## RESULTS

### Study cohort

The demographics and clinical characteristics of the study participants are summarized in Table 1. A total of 242 participants were recruited, consisting of manifesting carriers (PD-G2385R [*n*=57], PD-R1628P [*n*=61], PD-G2385R+R1628P [*n*=5]), iPD (*n*=61), and HC (*n*=58). There were no significant between-group differences in age, sex, body mass index, and comorbidities.

**Table 1:** Demographics and clinical characteristics of study participants.

|  | HC<br>(n=58) | iPD<br>(n=61) | PD-G2385R<br>(n=57) | PD-R1628P<br>(n=61) | PD-G2385R+R1628P<br>(n=5) | p-value |
| --- | --- | --- | --- | --- | --- | --- |
| **Age** | 67.3 ± 7.5 | 70.2 ± 9.3 | 69.5 ± 9.2 | 67.5 ± 8.5 | 70.4 ± 8.4 | 0.268 |
| **Sex** |  |  |  |  |  |  |
| % Male | 50.0 | 52.5 | 49.1 | 49.2 | 80.0 | 0.751 |
| **Body mass index (kg/m<sup>2</sup>)** | 24.2 [4.0] | 23.0 [8.6] | 23.0 [5.9] | 24.0 [6.9] | 23.0 [6.9] | 0.349 |
| **Comorbidities** |  |  |  |  |  |  |
| % Diabetes mellitus | 17.2 | 21.3 | 14.0 | 14.8 | 20.0 | 0.905 |
| % Hypertension | 44.8 | 44.3 | 49.1 | 34.4 | 20.0 | 0.756 |
| % Dyslipidemia | 56.9 | 47.5 | 31.6 | 39.3 | 20.0 | 0.195 |
| % Stroke | 3.5 | 9.8 | 3.5 | 1.6 | 20.0 | 0.304 |
| % Heart disease | 12.1 | 19.7 | 8.8 | 9.8 | 20.0 | 0.634 |
| % Kidney disease | 1.7 | 1.6 | 5.3 | 1.6 | 0.0 | 0.798 |
| % Cancer | 3.5 | 1.6 | 3.5 | 4.9 | 0.0 | 0.909 |
| **PD history** |  |  |  |  |  |  |
| Age at diagnosis (years) | NA | 62.1 ± 9.2 | 61.9 ± 9.8 | 60.0 ± 9.0 | 58.0 ± 7.3 | 0.488 |
| Disease duration (years) | NA | 8.0 [9.0] | 6.0 [7.0] | 6.0 [7.0] | 9.0 [2.0] | 0.645 |
| % With family history | NA | 18.0 | 24.6 | 21.3 | 80.0 | **0.009\*** |
| **PD severity** |  |  |  |  |  |  |
| Hoehn and Yahr stage<sup>a</sup> | NA | 2.0 [1.0] | 2.0 [1.0] | 2.0 [1.0] | 3.5 [1.5] | 0.304 |
| **MDS-UPDRS** |  |  |  |  |  |  |
| Part I score<sup>a</sup> | NA | 8.0 [10.2] | 8.0 [10.0] | 9.0 [7.5] | 7.0 [7.5] | 0.850 |
| Part II score<sup>a</sup> | NA | 12.0 [15.0] | 9.0 [10.0] | 11.0 [14.0] | 17.0 [20.0] | 0.258 |
| Part III score<sup>a</sup> | NA | 39.7 ± 14.6 | 45.3 ± 15.5 | 40.8 ± 13.6 | 58.5 ± 17.7 | **0.028\*** |
| Part IV score<sup>a</sup> | NA | 2.0 [6.0] | 2.0 [5.0] | 2.0 [6.0] | 1.5 [5.0] | 0.971 |
| Total score<sup>a</sup> | NA | 63.0 [41.0] | 66.5 [27.0] | 61.0 [38.5] | 93.0 [42.5] | 0.409 |
| **CISI-PD** |  |  |  |  |  |  |
| Motor signs<sup>a</sup> | NA | 3.0 [2.0] | 3.0 [1.0] | 3.0 [1.3] | 4.5 [1.5] | 0.259 |
| Disability<sup>a</sup> | NA | 3.0 [0.0] | 3.0 [1.0] | 3.0 [0.3] | 4.0 [2.3] | 0.353 |
| Motor complications<sup>a</sup> | NA | 1.0 [3.0] | 1.0 [2.0] | 1.0 [2.0] | 2.0 [2.3] | 0.694 |

**Table 1: Demographics and clinical characteristics of study participants (continued).**
|  | HC<br>(n=58) | iPD<br>(n=61) | PD-G2385R<br>(n=57) | PD-R1628P<br>(n=61) | PD-G2385R+R1628P<br>(n=5) | *p*-value |
| --- | --- | --- | --- | --- | --- | --- |
| Cognitive status<sup>a</sup> | NA | 2.0 [1.0] | 2.0 [2.0] | 2.0 [1.0] | 2.0 [1.0] | 0.483 |
| Total score<sup>a</sup> | NA | 9.0 [6.0] | 10.0 [6.0] | 10.0 [4.0] | 11.0 [1.0] | 0.571 |
| Motor subtypes |  |  |  |  |  | 0.387 |
| % TD | NA | 24.6 | 28.1 | 18.0 | 0.0 |  |
| % PIGD | NA | 65.6 | 57.9 | 65.6 | 100.0 |  |
| % Indeterminate | NA | 6.6 | 10.5 | 14.8 | 0.0 |  |
| % With MRC | NA | 60.7 | 63.2 | 59.0 | 60.0 | 0.975 |
| **Cognitive function** |  |  |  |  |  |  |
| MoCA score | 27.0 [3.0] | 25.0 [7.0] | 25.0 [6.3] | 26.0 [7.3] | 24.5 [2.0] | **0.003\*** |
| **PD treatment** |  |  |  |  |  |  |
| LEDD | NA | 450.0 [500.0] | 450.0 [400.0] | 450.0 [482.0] | 598.0 [295.0] | 0.936 |
| % With deep brain stimulation | NA | 3.3 | 12.3 | 8.2 | 20.0 | 0.244 |

For continuous variables, normally distributed data are reported as mean ± standard deviation, while non-normally distributed data are presented as median [interquartile range]. Categorical data are reported as %. *p*-value is obtained using ANOVA (normally distributed data), Kruskal-Wallis test (non-normally distributed data) or chi-square test (categorical data). Between-group differences with *p*-value<0.05 are considered significant (indicated by \*). <sup>a</sup>Patients on deep brain stimulation or apomorphine infusion excluded from analysis (*n*=16). Abbreviations: CISI-PD=Clinical Impression of Severity Index for Parkinson's Disease; HC=healthy controls; iPD=patients with idiopathic Parkinson's disease; LEDD=levodopa equivalent daily dose; MDS-UPDRS=International Parkinson and Movement Disorder Society-Unified Parkinson's Disease Rating Scale; MoCA=Montreal Cognitive Assessment; MRC=motor response complications; NA=not applicable; PD-G2385R=PD patients carrying the p.G2385R variant; PD-G2385R+R1628P=PD patients carrying both p.G2385R and p.R1628P; PD-R1628P=PD patients carrying p.R1628P; PIGD=postural instability-gait difficulty; TD=tremor dominant.

### Higher pRab10^Thr73^ phosphorylation levels in manifesting *LRRK2* p.G2385R and double-variant carriers

We observed a significant but modest increase in pRab10^Thr73^ phosphorylation in PD-G2385R (∼1.2-fold vs. HC; adjusted *p*=0.011) and a striking elevation in double-variant carriers (∼2.8-fold vs. HC; adjusted *p*=0.008) (Figure 1a). There were no significant differences between HC vs. iPD (adjusted *p*=0.073) or HC vs. PD-R1628P (adjusted *p*=0.159) in pRab10^Thr73^ phosphorylation levels. Compared to iPD, pRab10^Thr73^ phosphorylation was significantly higher in PD-G2385R (∼1.8-fold; adjusted *p*<0.001), PD-R1628P (∼1.6-fold; adjusted *p*=0.002), and especially in double-variant carriers (∼4.0-fold; adjusted *p*<0.001) (Supplementary Figure 2a). No significant differences were detected between PD-G2385R vs. PD-R1628P or between PD-G2385R vs. double-variant carriers. Total Rab10 levels were relatively consistent across all participants and did not differ significantly between groups (Figure 1b, Supplementary Figure 2b).

**Figure 1:**
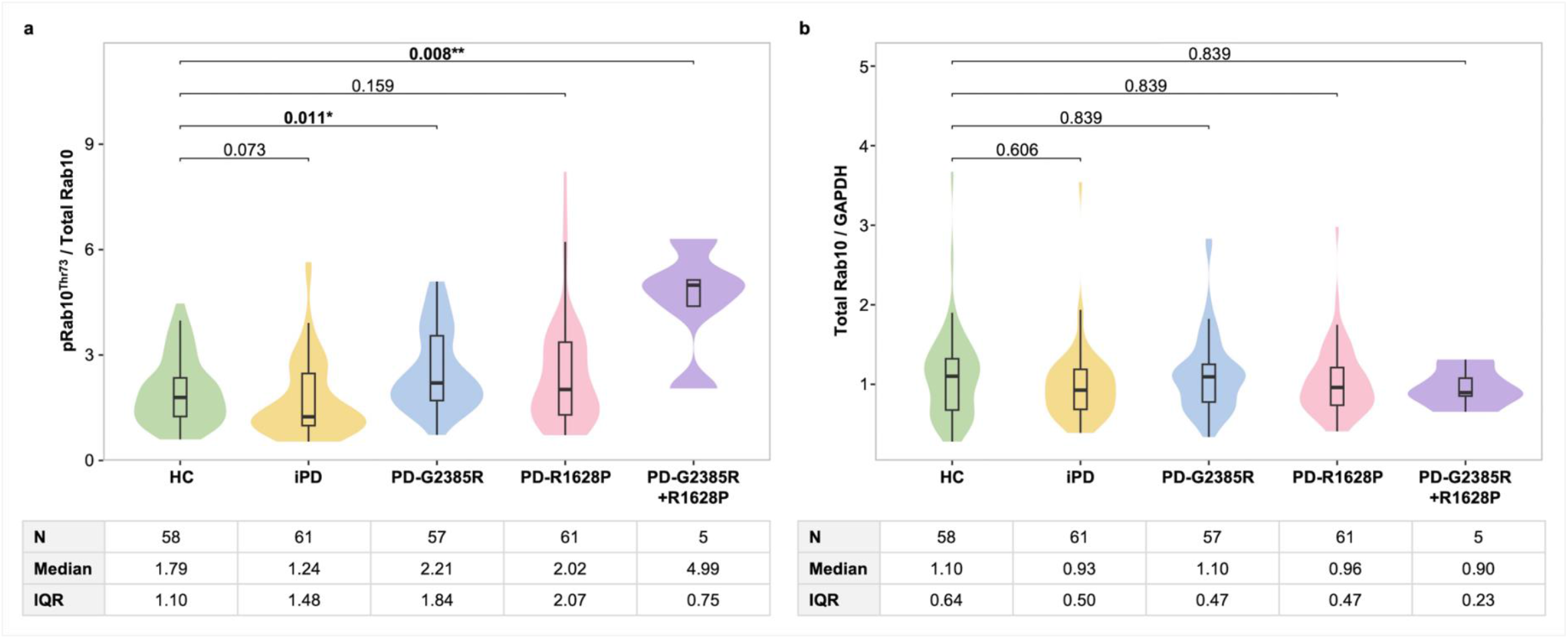
pRab10^Thr73^ phosphorylation in iPD, PD with *LRRK2* p.G2385R and/or p.R1628P variants, and controls. Monocyte lysates were analyzed by quantitative immunoblotting. Quantified immunoblotting data are presented as ratios of (**a**) pRab10^Thr73^/total Rab10 (Kruskal-Wallis test, *p*<0.001***) and (**b**) total Rab10/GAPDH (Kruskal-Wallis test, *p*=0.483), normalized to the average values obtained from an independent healthy control (experiment performed in duplicate). Pairwise comparisons between groups were conducted using the Kruskal-Wallis test with post hoc Dunn’s test, with *p*-values adjusted using the Benjamini-Hochberg method (adjusted *p*<0.05). The *p*-values for all pairwise comparisons are provided in Supplementary Figure 2. \**p*<0.05, \*\**p*<0.01, \*\*\**p*<0.001. Abbreviations: HC=healthy controls; iPD=patients with idiopathic Parkinson’s disease; IQR=interquartile range; PD-G2385R=PD patients carrying the p.G2385R variant; PD-G2385R+R1628P=PD patients carrying both p.G2385R and p.R1628P; PD-R1628P=PD patients carrying p.R1628P.

### Reduced pLRRK2^Ser935^ phosphorylation in manifesting *LRRK2* p.G2385R, p.R1628P, and double-variant carriers

For the analyses of pLRRK2^Ser935^ and total LRRK2 levels, 20 samples were excluded due to either unquantifiable LRRK2 bands (*n*=10) or total LRRK2/GAPDH ratio was <0.02 (*n*=10), leaving 222 samples for analysis. Compared with HC, pLRRK2^Ser935^ phosphorylation was significantly reduced in PD-G2385R (∼0.9-fold, adjusted *p*=0.007) and PD-R1628P (∼0.8-fold, adjusted *p*=0.002), with the lowest levels observed in the double-variant carriers (∼0.5-fold, adjusted *p*<0.001) (Figure 2a). Relative to iPD, pLRRK2^Ser935^ phosphorylation was significantly lower in PD-G2385R (adjusted *p*=0.021), PD-R1628P (adjusted *p*=0.006), and double-variant carriers (adjusted *p*<0.001) (Supplementary Figure 3a). Pairwise comparisons further demonstrated that double-variant carriers exhibited lower pLRRK2^Ser935^ phosphorylation than both PD-G2385R (adjusted *p*=0.006) and PD-R1628P (adjusted *p*=0.007), whereas levels were comparable between PD-G2385R and PD-R1628P (adjusted *p*=0.565). Total LRRK2 protein levels were consistent, with no significant differences observed across groups (Figure 2b, Supplementary Figure 3b). Notably, pLRRK2^Ser935^ phosphorylation correlated significantly with pRab10^Thr73^ phosphorylation (*r*_*s*_=-0.611, *p*<0.001).

**Figure 2:**
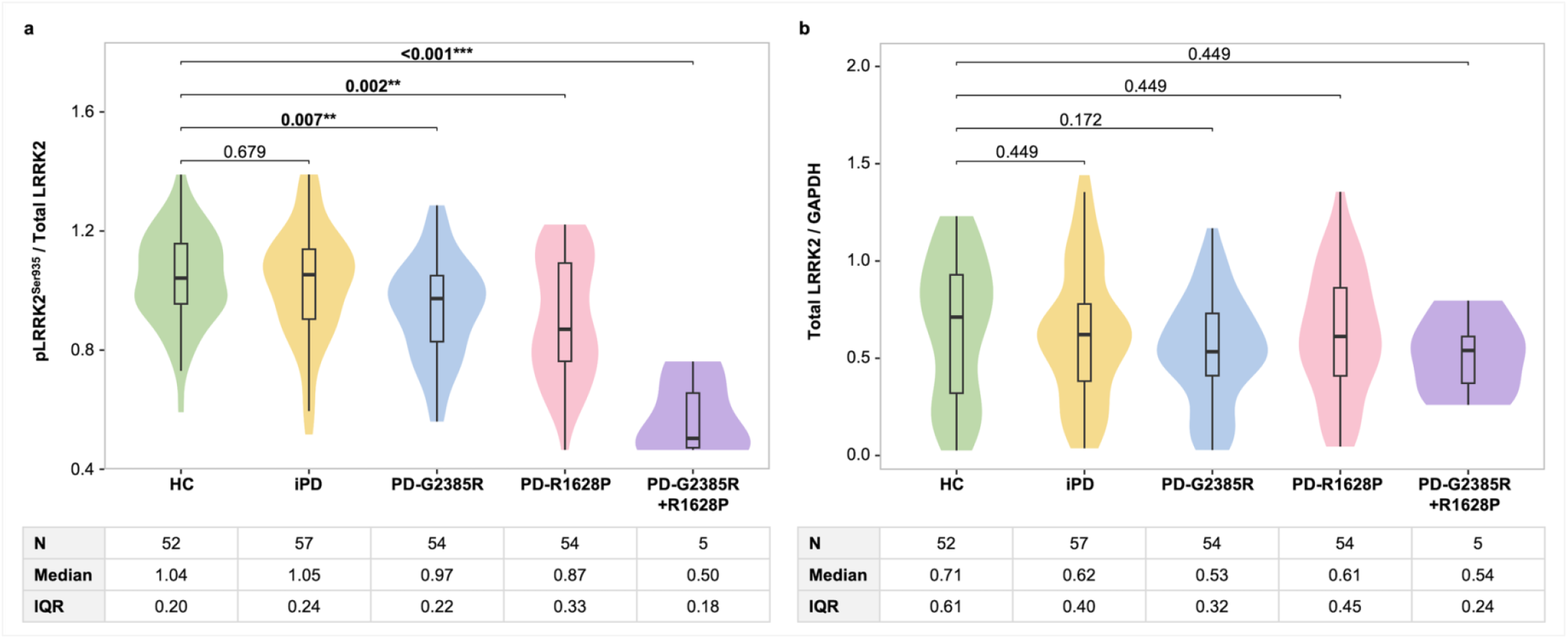
pLRRK2^Ser935^ phosphorylation in iPD, PD with *LRRK2* p.G2385R and/or p.R1628P variants, and controls. Monocyte lysates were analyzed by quantitative immunoblotting. Samples with faint or unquantifiable LRRK2 bands and total LRRK2/GAPDH values<0.02 were excluded from the analysis (*n*=20). Quantified immunoblotting data are presented as ratios of (**a**) pLRRK2^Ser935^/total LRRK2 (Kruskal-Wallis, *p*<0.001***) and (**b**) total LRRK2/GAPDH (Kruskal-Wallis, *p*<0.314), normalized to the average values obtained from an independent healthy control (experiment performed in duplicate). Pairwise comparisons between groups were conducted using the Kruskal-Wallis test with post hoc Dunn’s test, with *p*-values adjusted using the Benjamini-Hochberg method (adjusted *p*<0.05). The *p*-values for all pairwise comparisons are provided in Supplementary Figure 3. \**p*<0.05, \*\**p*<0.01, \*\*\**p*<0.001. Abbreviations: HC=healthy controls; iPD=patients with idiopathic Parkinson’s disease; IQR=interquartile range; PD-G2385R=PD patients carrying the p.G2385R variant; PD-G2385R+R1628P=PD patients carrying both p.G2385R and p.R1628P; PD-R1628P=PD patients carrying p.R1628P.

### Inter-individual variability in pRab10^Thr73^ and pLRRK2^Ser935^ phosphorylation

The distributions of individual pRab10^Thr73^ and pLRRK2^Ser935^ phosphorylation levels for all participants are displayed in Figure 3 and Supplementary Figures 4-5, alongside a pathogenic *LRRK2* p.R1067Q carrier (included as a positive control, this monocyte sample was processed in parallel with the study cohort using identical methodology^19^). For pRab10^Thr73^ phosphorylation, all double-variant carriers (*n*=5/5) exceeded the HC median, with levels comparable to that observed in the pathogenic p.R1067Q variant carrier. Meanwhile, we observed substantial inter-individual variation in other PD subgroups: the majority of *LRRK2* variant carriers (68.4% of PD-G2385R and 54.1% of PD-R1628P), as well as one-third (34.4%) of iPD, had levels above the median observed in HCs. A similar distribution was observed for pLRRK2^Ser935^ phosphorylation. When both measures were considered together, all double-variant carriers, 51.9% of PD-G2385R, 48.1% of PD-R1628P, and 28.1% of iPD exhibited the combined profile of elevated pRab10^Thr73^ and reduced pLRRK2^Ser935^ phosphorylation relative to HC medians.

**Figure 3:**
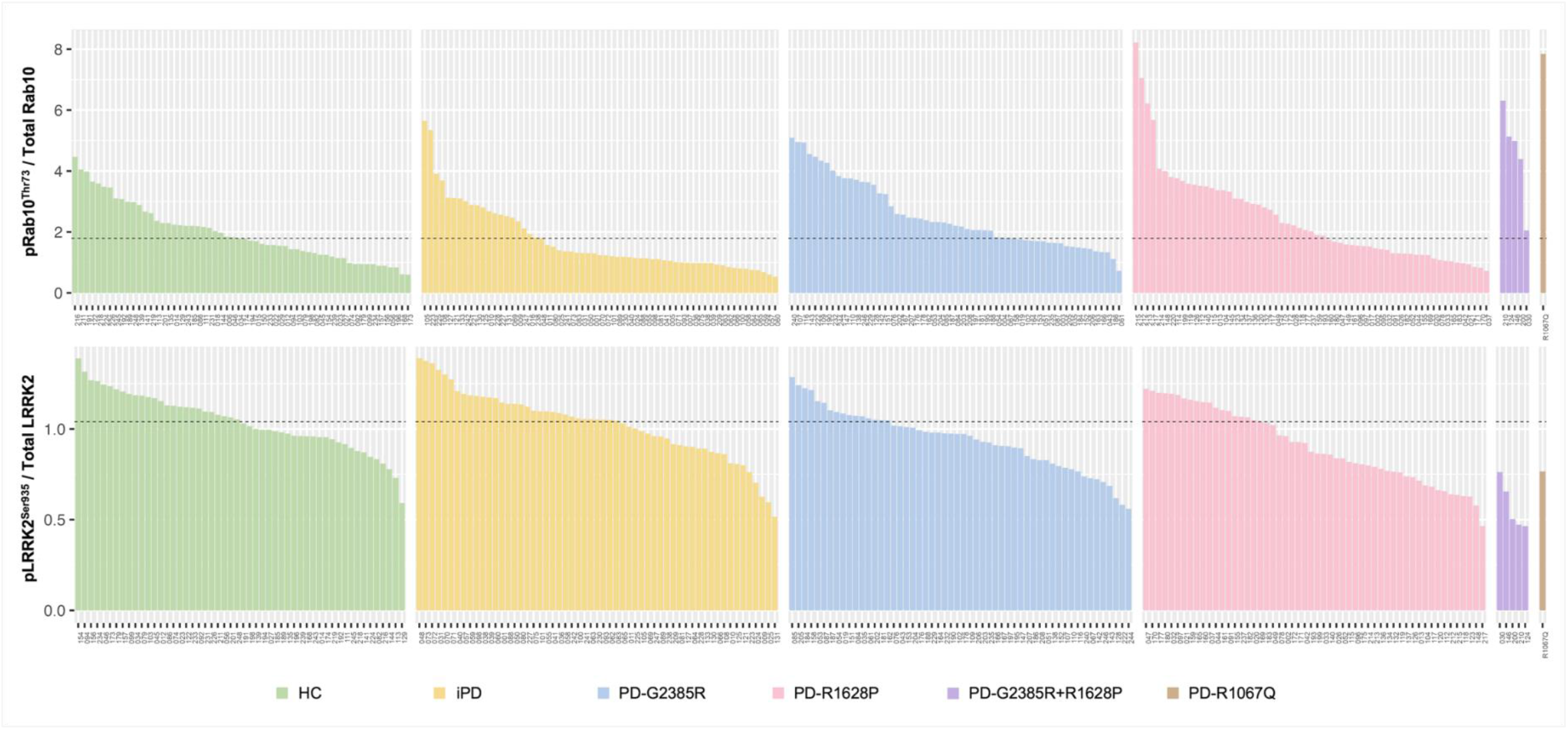
Distribution of pRab10^Thr73^ and pLRRK2^Ser935^ phosphorylation levels for each study participant. Dotted horizontal lines indicate the median values in the HC group (pRab10^Thr73^/total Rab10: median=1.79; pLRRK2^Ser935^/total LRRK2: median=1.04). A manifesting carrier of the pathogenic *LRRK2* p.R1067Q variant was included as a positive control. Inter-individual variation was observed in pRab10^Thr73^ and pLRRK2^Ser935^ phosphorylation. All double-variant carriers (*n*=5/5), 68.4% of PD-G2385R (*n*=39/57), 54.1% of PD-R1628P (*n*=33/61), and 34.4% of iPD (*n*=21/61) demonstrated pRab10^Thr73^ phosphorylation levels above the median of HC. Abbreviations: HC=healthy controls; iPD=patients with idiopathic Parkinson’s disease; PD-G2385R=PD patients carrying the p.G2385R variant; PD-G2385R+R1628P=PD patients carrying both p.G2385R and p.R1628P; PD-R1067Q=PD patient carrying p.R1067Q; PD-R1628P=PD patients carrying p.R1628P.

Importantly, none of the 232 of 242 participants who passed NBA quality control were found to carry other known pathogenic or likely pathogenic variants in *LRRK2, VPS35*, or *RAB32*, as listed in the Methods section above.

### Correlations with key clinical variables in PD

In the overall PD cohort, higher pRab10^Thr73^ phosphorylation levels were significantly associated with more favorable clinical outcomes. Specifically, higher pRab10^Thr73^ phosphorylation correlated with reduced motor severity (lower CISI-PD motor scores [*r*_*s*_=-0.152, *p*=0.0495]), less disability (lower CISI-PD disability scores [*r*_*s*_=-0.160, *p*=0.039] and lower MDS-UPDRS Part II scores [*r*_*s*_=-0.155, *p*=0.045]), reduced non-motor symptom burden (lower MDS-UPDRS Part I scores [*r*_*s*_=-0.156, *p*=0.044]), and better cognition (higher MoCA scores [*r*_*s*_=0.157, *p*=0.042]) (Figure 4a). Subgroup analyses revealed consistent trends: in the iPD subgroup, higher pRab10^Thr73^ phosphorylation were significantly associated with lower CISI-PD disability scores (*r*_*s*_=-0.283, *p*=0.038), while in the PD-R1628P subgroup, they were associated with lower CISI-PD motor sign scores (*r*_*s*_=-0.276, *p*=0.039). After covariate adjustment (Supplementary Table 2), only one association remained significant: higher pRab10^Thr73^ phosphorylation correlated with better MoCA score in the overall PD cohort (*p*=0.020; Figure 4b).

**Figure 4:**
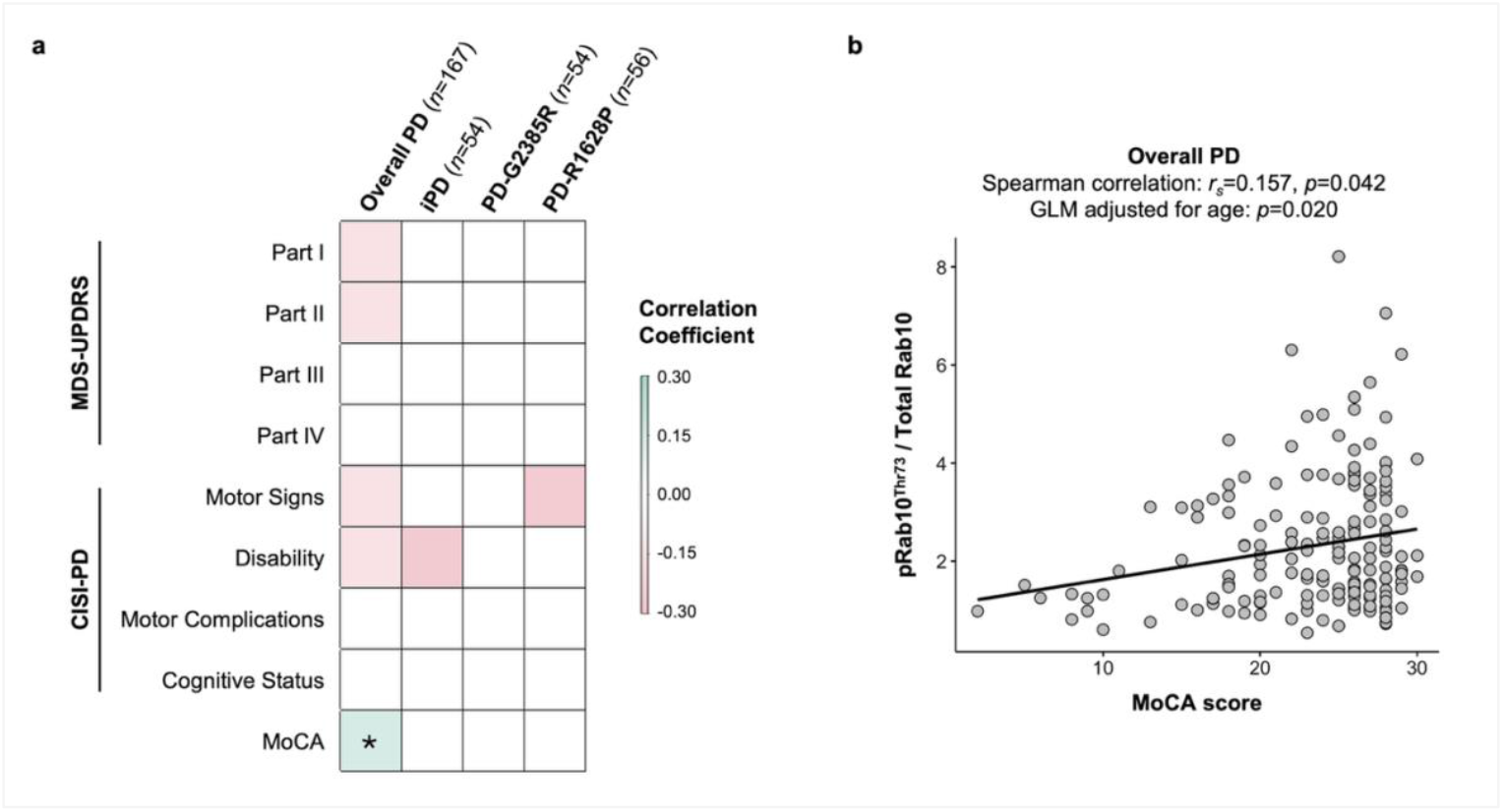
Correlations of pRab10^Thr73^ phosphorylation levels with key clinical variables in PD. (**a**) Correlogram showing the correlations of pRab10^Thr73^/total Rab10 levels with various clinical variables in the overall PD group, and within the iPD, PD-G2385R, and PD-R1628P subgroups (Spearman correlation, *p*<0.05). Green indicates significant positive correlations, red indicates significant negative correlations, with increasing darkness representing stronger associations. (**b**) Scatter plot showing the positive correlation between pRab10^Thr73^/total Rab10 and MoCA scores in the overall PD group, which remained significant after adjustment for age using a generalized linear model (GLM). Higher MoCA scores reflect better cognitive status, whereas higher MDS-UPDRS and CISI-PD scores indicate worse clinical status. Abbreviations: CISI-PD=Clinical Impression of Severity Index for Parkinson’s Disease; GLM=generalized linear model; iPD=patients with idiopathic Parkinson’s disease; MDS-UPDRS=International Parkinson and Movement Disorder Society-Unified Parkinson’s Disease Rating Scale; MoCA=Montreal Cognitive Assessment; PD-G2385R=PD patients carrying the p.G2385R variant; PD-R1628P=PD patients carrying p.R1628P.

For pLRRK2^Ser935^ phosphorylation, no significant correlations with clinical variables were observed in the overall PD cohort or in the PD-G2385R and PD-R1628P subgroups (Supplementary Table 3). However, in the iPD subgroup, higher pLRRK2^Ser935^ phosphorylation were associated with worse MDS-UPDRS Part III (*r*_*s*_=0.286, *p*=0.044), CISI-PD motor signs (*r*_*s*_=0.300, *p*=0.035) and CISI-PD disability scores (*r*_*s*_=0.347, *p*=0.014). These associations were not significant after adjustment for age and disease duration.

## DISCUSSION

Using a robust multiplexed immunoblotting assay, we demonstrated significant alterations in LRRK2 kinase pathway activity in PD patients carrying Asian-prevalent variants. pRab10^Thr73^ phosphorylation was modestly elevated in PD-G2385R (∼1.2-fold vs. controls) and markedly increased in double-variant carriers (∼2.8-fold vs. controls), suggesting an additive or even synergistic effect of these variants. Levels were also higher in all *LRRK2* variant subgroups compared with iPD. In parallel, pLRRK2^Ser935^ phosphorylation was consistently reduced in PD-G2385R, PD-R1628P, and double-variant manifesting carriers than HCs, suggesting that LRRK2 adopts a more active conformation in these patient samples. All double-variant carriers and the majority of single-variant carriers exhibited elevated pRab10^Thr73^ phosphorylation with corresponding reduction in pLRRK2^Ser935^ phosphorylation, when compared to the median levels in HC. The inter-individual variability in kinase activity among single-variant carriers highlights the influence of additional genetic or environmental modifiers and underscores the need for biochemical stratification beyond genotyping in future LRRK2-targeted trials. Intriguingly, approximately one-third of iPD also demonstrated high kinase activity despite the absence of known LRRK2-activating exonic variants, suggesting that LRRK2 hyperactivation is not limited to genetically-predisposed cases, thus highlighting the potential opportunities for LRRK2-targeted interventions even in ‘idiopathic’ cases. Taken together, this work represents the largest human study of LRRK2 phosphorylation assays and biochemical investigation of Asian-prevalent *LRRK2* variant carriers to date, extending evidence from predominantly cell-based models to a large, genotype-stratified patient cohort.

LRRK2 is highly expressed in immune cells,^37^ and plays a key role in regulating autophagy, lysosomal function, and membrane trafficking.^6^ Among its substrates, Rab GTPases such as Rab10 are particularly relevant, as their phosphorylation disrupts vesicle transport, docking, and cilia formation—processes implicated in PD pathogenesis.^6,38,39^ Although only a small fraction of Rab proteins are phosphorylated under steady-state conditions, even modest increases can trigger abnormal interactions with effector proteins, such as RILPL1 and RILPL2, leading to measurable physiological consequences.^40^ In our study, PD-G2385R exhibited a relatively subtle ∼1.2-fold increase in pRab10^Thr73^ phosphorylation compared with controls, consistent with the concept that even small shifts in Rab10 phosphorylation may be biologically meaningful. In contrast, the ∼2.8-fold elevation observed in double-variant carriers—and in a pathogenic p.R1067Q carrier processed in parallel—represents a more pronounced increase in kinase activity. This aligns with prior work using a similar assay, which reported a ∼4.5-fold increase in pRab10^Thr73^ phosphorylation in neutrophils from carriers of the pathogenic p.R1441G variant,^17^ which has been reported to have high penetrance (>80% at 80 years).^41-43^ Collectively, these findings suggest a spectrum of LRRK2-driven Rab10 phosphorylation, ranging from subtle changes in risk variants (e.g., p.G2385R) to more robust elevations in double-variant or pathogenic carriers. We also observed a similar pattern in pLRRK2^Ser935^ phosphorylation, which was more markedly reduced in double-variant carriers than risk variants. This gradient raises the important possibility that the degree of kinase inhibition required for therapeutic efficacy may differ across patient subgroups, an outstanding question in the field.^44^ Importantly, our ability to capture this dose-dependent effect on Rab10 phosphorylation using a relatively straightforward immunoblotting assay, together with prior work showing strong concordance with mass spectrometry measurements,^17^ supports the utility of this approach, particularly in research or clinical settings where mass spectrometry is not readily available.

Multiple studies have reported conflicting effects of the Asian-prevalent p.G2385R and p.R1628P variants on LRRK2 kinase activity. Structurally, these variants were hypothesized to disrupt critical protein-protein interactions involved in regulating LRRK2 function via alterations of the WD40 domain dimerization interface and the COR_A_-COR_B_ domain interactions, respectively.^13,28^ p.G2385R has been described as a partial loss-of-function mutation in some *in vitro* assays, with reduced autophosphorylation or substrate phosphorylation,^23,24^ while other cellular assays reported normal^25^ or elevated^13,26-28^ kinase activity. Much less is known about the mechanisms of p.R1628P. Two *in vitro* kinase assays have demonstrated increased kinase activity in p.R1628P, possibly indirectly via cyclin-dependent kinase 5 phosphorylation of LRRK2 at S1627,^29^ while one showed comparable kinase activity to wild-type.^13^ These disparate findings likely reflect methodological differences—early in vitro studies often used isolated protein and artificial substrates, whereas more recent work has leveraged cellular and physiological Rab phosphorylation assays—and highlight that genetic background (e.g., protective variants, such as p.R1398H or p.N551K) or environmental toxins (e.g., MPP+), as well as gene-environment interactions, can potentially alter the effects of these risk alleles.^27,29^

In light of these mixed results, our study provides compelling *in vivo* evidence that p.G2385R can enhance LRRK2 kinase activity in human-derived monocytes, consistent with previous reports of increased pRab10^Thr73^ phosphorylation levels in HEK293 cellular assay^13^ and in post-mortem brain sample from a patient with p.G2385R.^45^ In PD-R1628P patients, we found reduced pLRRK2^Ser935^ phosphorylation without a significant increase in pRab10^Thr73^ phosphorylation, mirroring prior findings in HEK293 cellular assay.^13^ However, the more than two-fold increase in kinase activity among double-variant carriers suggests that p.R1628P can contribute to increased kinase activity, possibly through synergistic or context-dependent mechanisms, for example, when combined with other kinase activating variants. Notably, we observed substantial inter-individual variability in kinase readouts: some single-variant carriers (and even certain non-carriers) displayed robust hyperactivation, whereas others were closer to baseline seen in HC. None of the individuals in our cohort harbored other known kinase-activating variants, again highlighting the role of other genetic or environmental modifiers of LRRK2 kinase activity. This heterogeneity underscores the need for biochemical stratification beyond genotyping, echoing similar variability seen recently in cerebrospinal fluid α-synuclein seed amplification assay (CSF SAA) data, where only two-thirds of *LRRK2* p.G2019S carriers tested positive,^46^ despite the fact that synucleinopathy has long been considered a hallmark— or even defining—feature of PD.^47-49^ Emerging evidence that genetic variation in molecular regulators, such as RAB29, influences CSF SAA status,^50^ further highlights the importance of investigating modifiers of LRRK2 activity and of downstream pathways impacted by *LRRK2* variation.

Besides the novel insights on p.G2385R and p.R1628P carriers, our finding of high pRab10^Thr73^ phosphorylation in approximately one-third of iPD deserve attention. This observation aligns with genomic data linking PD risk to non-coding variants at the *LRRK2* locus, and with prior studies demonstrating increased LRRK2 kinase activity in nigrostriatal dopamine neurons of iPD.^44,51^ Other human studies support this broader relevance: flow cytometry-based assays have shown increased LRRK2 and Rab10 phosphorylation in peripheral blood mononuclear cells of iPD,^52^ and urinary extracellular vesicle studies have reported increased LRRK2 autophosphorylation and Rab phosphorylation in subsets of iPD,^7,53^ although replication has been inconsistent. Together, these findings support the concept that wild-type LRRK2 can be hyperactivated in iPD, and that LRRK2 kinase inhibition could potentially be deployed in this (much larger) group of patients.^54^

Intriguingly, we found that higher pRab10^Thr73^ phosphorylation levels correlated with better clinical outcomes—lower motor and disability burden, as well as better cognition— among PD patients in our overall cohort. In our previous deep phenotypic characterization study, both p.G2385R and p.R1628P carriers demonstrated slower disease progression compared to non-carriers.^21^ Additionally, p.R1628P carriers in our cohort were found to have significantly lower rates of dementia,^21^ while a meta-analysis reported better cognitive scores among p.G2385R carriers.^55^ Collectively, a picture seems to be emerging of a slightly more ‘benign’ progression of disease with the Asian-prevalent *LRRK2* risk variants compared with iPD, akin to what has been observed with p.G2019S.^21,56,57^ More recently, *LRRK2*-parkinsonism patients without evidence of α-synuclein aggregates were found to have less severe motor manifestations and decline, compared to those with α-synuclein aggregates.^50^ Although seemingly counter-intuitive, these positive associations hint at a complex interplay between LRRK2 signaling, α-synuclein aggregation, and disease manifestations, warranting further investigation into whether elevated kinase activity could reflect compensatory mechanisms or patient subgroups with distinct disease trajectories. This underscores the need for further investigation with larger sample sizes to validate and elucidate these observations, especially as several LRRK2 kinase inhibitors are already in clinical trials.^2,4,58^ While significant progress has been made in understanding LRRK2 protein kinase biology over the past decade, these findings highlight the urgent need for a more comprehensive understanding of the heterogeneous mechanisms underlying *LRRK2*-PD development and progression, which remain unresolved.

Our study has several limitations. First, although this represents the largest biochemical analysis of Asian-prevalent *LRRK2* variant carriers to date, the number of double-variant carriers was small, limiting the generalizability of conclusions about *LRRK2* genetic burden and kinase hyperactivation. Second, phosphorylation measurements were derived from peripheral monocytes, which may not fully reflect LRRK2 kinase activity in the brain. Nevertheless, prior studies have shown increased pRab10^Thr73^ phosphorylation in post-mortem brain tissue of iPD and a p.G2385R carrier,^45,51^ although reliable central LRRK2 target engagement markers, such as positron emission tomography tracer and cerebrospinal fluid assays, remain under development.^44^ Notably, monocytes have lower protease activity than neutrophils and are less prone to LRRK2 degradation during processing.^11^ Third, despite using a robust multiplexed immunoblotting method, variability in sample quality and assay performance may have contributed to the observed inter-individual heterogeneity. Longitudinal sampling will be required to further examine assay stability over time. Integrating both markers—elevated pRab10^Thr73^ phosphorylation together with reduced pLRRK2^Ser935^ phosphorylation—may provide a more reliable and cross-validated measure of LRRK2 activity, helping to overcome the limitations of each individual marker. Lastly, while none of the individuals were identified to have known LRRK2 kinase-activating variants, we cannot exclude the contribution of other rare or polygenic modifiers, or environmental exposures, which remain to be systematically explored. Replication in larger, independent cohorts, alongside integration with genetic, environmental, and multi-omics data, will be essential to validate and extend these findings.

## CONCLUSION

In summary, our study provides the largest *in vivo* biochemical assessment of LRRK2 kinase activity in Asian-prevalent variant carriers, demonstrating that the p.G2385R variant, and potentially p.R1628P, can enhance Rab10 phosphorylation in human-derived monocytes, with the most pronounced effects observed in double-variant carriers. We further show that a considerable proportion of iPD also exhibit elevated LRRK2 activity, underscoring the broader relevance of LRRK2 signaling beyond patients harboring coding mutations. The observed inter-individual variability in kinase activity, independent of known pathogenic variants, highlights the influence of additional genetic and environmental modifiers, and reinforces the need for precision biochemical stratification beyond genotyping in future LRRK2 inhibitor trials. Importantly, these findings emphasize the global relevance of biomarker-driven approaches in developing disease-modifying therapeutics in PD, and the urgent need to expand research on LRRK2 biology in under-represented populations.

## Data Availability

The data supporting the findings of this study are available from the corresponding author upon reasonable request. These data are not publicly available due to privacy and ethical restrictions.

## ACKNOWLEDGEMENTS

The authors gratefully acknowledge funding from the Michael J. Fox Foundation for Parkinson’s Research (MJFF), which supported a collaborative research effort between the Universiti Malaya, Malaysia, and the University of Dundee, United Kingdom, for the discovery of Asian LRRK2 biomarkers (MJFF-010188, MJFF-021041, MJFF-022659, awarded to AHT, SYL, and ES). The work was also supported by the Universiti Malaya Parkinson’s Disease and Movement Disorders Research Program (PV035-207, awarded to AHT and SYL). We would also like to thank Samantha Hutten and Rebecca Ouellette from MJFF for their kind assistance and support in the project and grant management. NBA data used in the preparation of this article were obtained from Global Parkinson’s Genetics Program (GP2). GP2 is funded by the Aligning Science Across Parkinson’s (ASAP) initiative and implemented by The Michael J. Fox Foundation for Parkinson’s Research (https://gp2.org). For a complete list of GP2 members see https://gp2.org. We sincerely thank all participants for their invaluable contributions, without which this research would not have been possible.

