## Supplementary for "Precision Stratification: Kinase Assay in Asian *LRRK2* Risk Carriers and Idiopathic Parkinson Disease"

**Supplementary Materials**


| | **Page No.** |
|---|---|
| **Supplementary Methods** | **2** |
| **Supplementary Tables** | **3 - 5** |
| Supplementary Table 1 | 3 |
| Supplementary Table 2 | 4 |
| Supplementary Table 3 | 5 |
| **Supplementary Figures** | **6 - 10** |
| Supplementary Figure 1 | 6 |
| Supplementary Figure 2 | 7 |
| Supplementary Figure 3 | 8 |
| Supplementary Figure 4 | 9 |
| Supplementary Figure 5 | 10 |

### Supplementary Methods

#### Human monocyte isolation from peripheral fresh blood

Twenty mL of fresh blood was collected from the subjects using EDTA blood tubes. Within an hour of sample collection, monocytes were isolated from the blood samples using the EasySep™ Direct Human Monocyte Isolation Kit (STEMCELL Technologies, Vancouver, Canada, Cat#19669), according to a standardized protocol, as previously described.[1] Monocyte lysates were snap-frozen and stored at -80°C until further processing.

Before immunoblotting, the protein concentration of each sample was assessed using the bicinchoninic acid (BCA) assay (Pierce™ BCA Protein Assay Kits, Thermo Fisher Scientific™, Massachusetts, USA). After determining the protein concentration, samples were diluted with lysis buffer and NuPAGE™ lithium dodecyl sulfate sample buffer (4X) containing β-mercaptoethanol to achieve a final concentration of 10 μg/μL. The diluted samples were heated at 96°C for 10 minutes before immunoblotting.

#### Multiplexed quantitative immunoblotting for LRRK2 kinase pathway activation

Multiplexed quantitative immunoblotting was performed for $pRab10^{Thr73}$, $pLRRK2^{Ser935}$, total Rab10, total LRRK2, and GAPDH, as previously described.[2,3] Briefly, 2 μL of diluted samples were loaded into each well of NuPAGE 4-12% Bis-Tris Midi Gels (Thermo Fisher Scientific™, Massachusetts, USA) and electrophoresed at 90 V for 15 minutes followed by 150 V for 60-75 minutes. Samples were run in duplicate. Proteins were then transferred onto the Amersham™ Protran® 0.45 μm nitrocellulose membrane (Cytiva, Massachusetts, USA) via electrophoresis at 90 V for 90 minutes on ice in the transfer buffer. Membranes were blocked for 1 hour at room temperature with blocking solution (skimmed milk powder dissolved in TBS-T), washed with TBS-T, and incubated with primary antibodies at 4°C overnight. After three 5-minute washes with TBS-T, membranes were incubated with secondary antibodies for one hour at room temperature in the dark. This was followed by three 10-minute washes in TBS-T. Protein bands were acquired via near-infrared fluorescent detection using the Odyssey® CLx imaging system (LI-COR Biosciences, Nebraska, USA) and quantified using Image Studio Lite (version 5.2.5). The list of primary and secondary antibodies used in this assay is provided in Supplementary Table 1, and representative immunoblots are shown in Supplementary Figure 1.

**Supplementary Table 1: List of antibodies used in the immunoblotting assay.**

| **Epitope and host** | **Source of reference** | **Catalogue no.** |
|---|---|---|
| Primary antibodies | | |
| Anti-Rab10 (phospho T73) antibody *(rabbit monoclonal)* | Abcam | MJF-21-108-10 (ab230261) |
| Anti-Rab10 antibody *(mouse monoclonal)* | Nanotools | 0680–100/Rab10-605B11 |
| Anti-LRRK2 (phospho S935) antibody *(rabbit monoclonal)* | MRC PPU Reagents and Services | UDD2 |
| Anti-LRRK2 C-terminal antibody *(mouse monoclonal)* | Neuromab | 75-253 |
| Anti-GAPDH antibody *(mouse monoclonal)* | Santa Cruz Biotechnology | sc-32233 |
| Secondary antibodies | | |
| IRDye® 680LT Goat anti-Mouse | LICORbio | 926-68020 |
| IRDye® 800CW Goat anti-Rabbit | LICORbio | 926-32211 |

**Supplementary Table 2: Correlations of pRab10$^{Thr73}$ phosphorylation with key clinical variables overall and in *LRRK2*-stratified PD subgroups.**

| Clinical Variable | Spearman's rho | Spearman *p*-value | GLM *p*-value[a] |
|---|---|---|---|
| Overall PD cohort (*n*=167) | | | |
| MDS-UPDRS Part I | -0.156 | **0.044*** | 0.159 |
| MDS-UPDRS Part II | -0.155 | **0.045*** | 0.406 |
| MDS-UPDRS Part III | -0.046 | 0.558 | 0.422 |
| MDS-UPDRS Part IV | -0.087 | 0.262 | 0.234 |
| CISI-PD Motor Signs | -0.152 | **0.050*** | 0.928 |
| CISI-PD Disability | -0.160 | **0.039*** | 0.731 |
| CISI-PD Motor Complications | -0.141 | 0.068 | 0.103 |
| CISI-PD Cognitive Status | -0.046 | 0.558 | 0.804 |
| MoCA | 0.157 | **0.042*** | **0.020*** |
| iPD (*n*=54) | | | |
| MDS-UPDRS Part I | -0.194 | 0.160 | 0.766 |
| MDS-UPDRS Part II | -0.172 | 0.214 | 0.990 |
| MDS-UPDRS Part III | -0.154 | 0.267 | 0.519 |
| MDS-UPDRS Part IV | -0.157 | 0.258 | **0.048*** |
| CISI-PD Motor Signs | -0.266 | 0.052 | 0.813 |
| CISI-PD Disability | -0.283 | **0.038*** | 0.624 |
| CISI-PD Motor Complications | -0.246 | 0.073 | 0.098 |
| CISI-PD Cognitive Status | -0.114 | 0.414 | 0.975 |
| MoCA | 0.253 | 0.065 | 0.747 |
| PD-G2385R (*n*=54) | | | |
| MDS-UPDRS Part I | -0.065 | 0.640 | 0.937 |
| MDS-UPDRS Part II | -0.175 | 0.207 | 0.787 |
| MDS-UPDRS Part III | -0.116 | 0.404 | 0.611 |
| MDS-UPDRS Part IV | -0.008 | 0.957 | 0.807 |
| CISI-PD Motor Signs | -0.059 | 0.670 | 0.449 |
| CISI-PD Disability | -0.209 | 0.129 | 0.456 |
| CISI-PD Motor Complications | 0.037 | 0.792 | 0.421 |
| CISI-PD Cognitive Status | -0.239 | 0.081 | 0.054 |
| MoCA | 0.185 | 0.181 | 0.057 |
| PD-R1628P (*n*=56) | | | |
| MDS-UPDRS Part I | -0.153 | 0.260 | 0.273 |
| MDS-UPDRS Part II | -0.057 | 0.675 | 0.710 |
| MDS-UPDRS Part III | -0.091 | 0.505 | 0.461 |
| MDS-UPDRS Part IV | -0.021 | 0.881 | 0.435 |
| CISI-PD Motor Signs | -0.276 | **0.039*** | 0.071 |
| CISI-PD Disability | -0.194 | 0.151 | 0.276 |
| CISI-PD Motor Complications | -0.072 | 0.596 | 0.261 |
| CISI-PD Cognitive Status | 0.097 | 0.477 | 0.519 |
| MoCA | 0.080 | 0.557 | 0.305 |

[a]*p*-values derived from generalized linear model (GLM) controlling for covariates. Analyses of MDS-UPDRS Parts I-III, CISI-PD Motor Signs, and CISI-PD Disability were adjusted for age and disease duration; analyses of MDS-UPDRS Part IV and CISI-PD Motor Complications were adjusted for age at diagnosis and disease duration; analyses of MoCA and CISI-PD Cognitive Status were adjusted for age. **p*-values<0.05 were considered statistically significant. Abbreviations: GLM=generalized linear model; iPD=patients with idiopathic Parkinson's disease; PD-G2385R=PD patients carrying the p.G2385R variant; PD-R1628P=PD patients carrying p.R1628P.

**Supplementary Table 3: Correlations of pLRRK2$^{Ser935}$ phosphorylation with key clinical variables overall, and in *LRRK2*-stratified PD subgroups.**

| **Clinical Variable** | **Spearman's rho** | **Spearman *p*-value** | **GLM *p*-value**[a] |
|---|---|---|---|
| Overall PD cohort (*n*=153) | | | |
| MDS-UPDRS Part I | 0.018 | 0.824 | 0.950 |
| MDS-UPDRS Part II | 0.092 | 0.259 | 0.595 |
| MDS-UPDRS Part III | 0.011 | 0.892 | 0.652 |
| MDS-UPDRS Part IV | 0.076 | 0.349 | 0.432 |
| CISI-PD Motor Signs | 0.086 | 0.292 | 0.966 |
| CISI-PD Disability | 0.059 | 0.469 | 0.693 |
| CISI-PD Motor Complications | 0.092 | 0.257 | 0.713 |
| CISI-PD Cognitive Status | -0.006 | 0.942 | 0.822 |
| MoCA | -0.016 | 0.844 | 0.369 |
| iPD (*n*=50) | | | |
| MDS-UPDRS Part I | 0.008 | 0.955 | 0.138 |
| MDS-UPDRS Part II | 0.259 | 0.069 | 0.566 |
| MDS-UPDRS Part III | 0.286 | **0.044*** | 0.803 |
| MDS-UPDRS Part IV | 0.237 | 0.097 | 0.707 |
| CISI-PD Motor Signs | 0.300 | **0.035*** | 0.534 |
| CISI-PD Disability | 0.347 | **0.014*** | 0.640 |
| CISI-PD Motor Complications | 0.259 | 0.069 | 0.872 |
| CISI-PD Cognitive Status | 0.136 | 0.345 | 0.607 |
| MoCA | -0.149 | 0.302 | 0.437 |
| PD-G2385R (*n*=51) | | | |
| MDS-UPDRS Part I | -0.097 | 0.499 | 0.646 |
| MDS-UPDRS Part II | -0.132 | 0.356 | 0.592 |
| MDS-UPDRS Part III | -0.013 | 0.925 | 0.844 |
| MDS-UPDRS Part IV | 0.089 | 0.534 | 0.476 |
| CISI-PD Motor Signs | -0.080 | 0.575 | 0.520 |
| CISI-PD Disability | -0.091 | 0.526 | 0.487 |
| CISI-PD Motor Complications | 0.051 | 0.720 | 0.990 |
| CISI-PD Cognitive Status | 0.012 | 0.934 | 0.968 |
| MoCA | 0.050 | 0.728 | 0.831 |
| PD-R1628P (*n*=49) | | | |
| MDS-UPDRS Part I | 0.110 | 0.453 | 0.403 |
| MDS-UPDRS Part II | 0.089 | 0.545 | 0.448 |
| MDS-UPDRS Part III | -0.090 | 0.539 | 0.895 |
| MDS-UPDRS Part IV | -0.104 | 0.477 | 0.788 |
| CISI-PD Motor Signs | 0.135 | 0.355 | 0.168 |
| CISI-PD Disability | 0.046 | 0.752 | 0.688 |
| CISI-PD Motor Complications | -0.100 | 0.496 | 0.948 |
| CISI-PD Cognitive Status | -0.134 | 0.359 | 0.562 |
| MoCA | 0.063 | 0.669 | 0.929 |

[a]*p*-values derived from generalized linear model (GLM) controlling for covariates. Analyses of MDS-UPDRS Parts I-III, CISI-PD Motor Signs, and CISI-PD Disability were adjusted for age and disease duration; analyses of MDS-UPDRS Part IV and CISI-PD Motor Complications were adjusted for age at diagnosis and disease duration; analyses of CISI-PD Cognitive Status and MoCA were adjusted for age. **p*-values<0.05 were considered statistically significant. Abbreviations: GLM=generalized linear model; iPD=patients with idiopathic Parkinson's disease; PD-G2385R=PD patients carrying the p.G2385R variant; PD-R1628P=PD patients carrying p.R1628P.

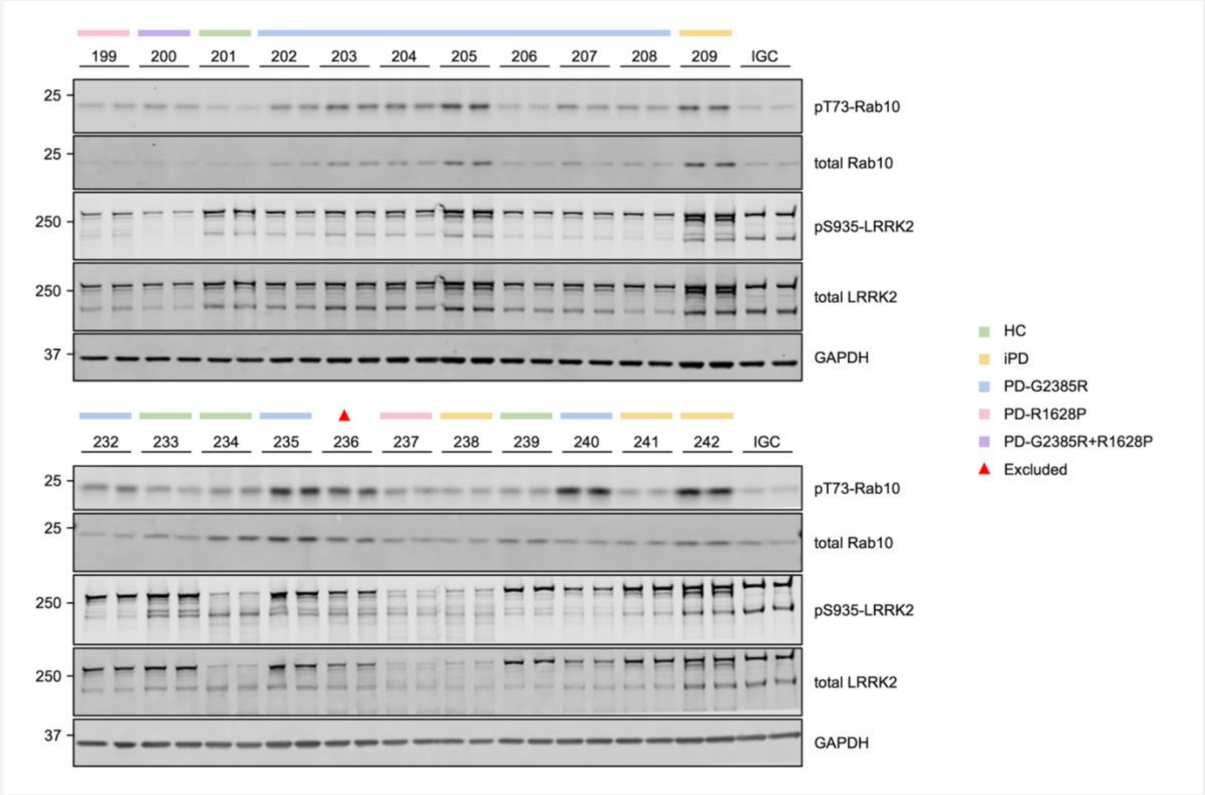


**Supplementary Figure 1: Representative immunoblot analysis of peripheral blood monocyte samples.**

Monocytes were isolated from fresh peripheral blood, and 20 µg of whole-cell extracts were loaded in duplicates for quantitative immunoblot analysis using the antibodies listed in Supplementary Table 1. Blots were developed using the Odyssey CLx Western Blot Imaging System. Phosphorylated Rab10 (pT73-Rab10) and total Rab10, as well as phosphorylated LRRK2 (pS935-LRRK2) and total LRRK2, were multiplexed. An internal standard ('IGC') was included on every gel to enable comparison across different gels. Abbreviations: HC=healthy controls; iPD=patients with idiopathic Parkinson's disease; PD-G2385R=PD patients carrying the p.G2385R variant; PD-G2385R+R1628P=PD patient carrying both p.G2385R and p.R1628P; PD-R1628P=PD patients carrying p.R1628P.

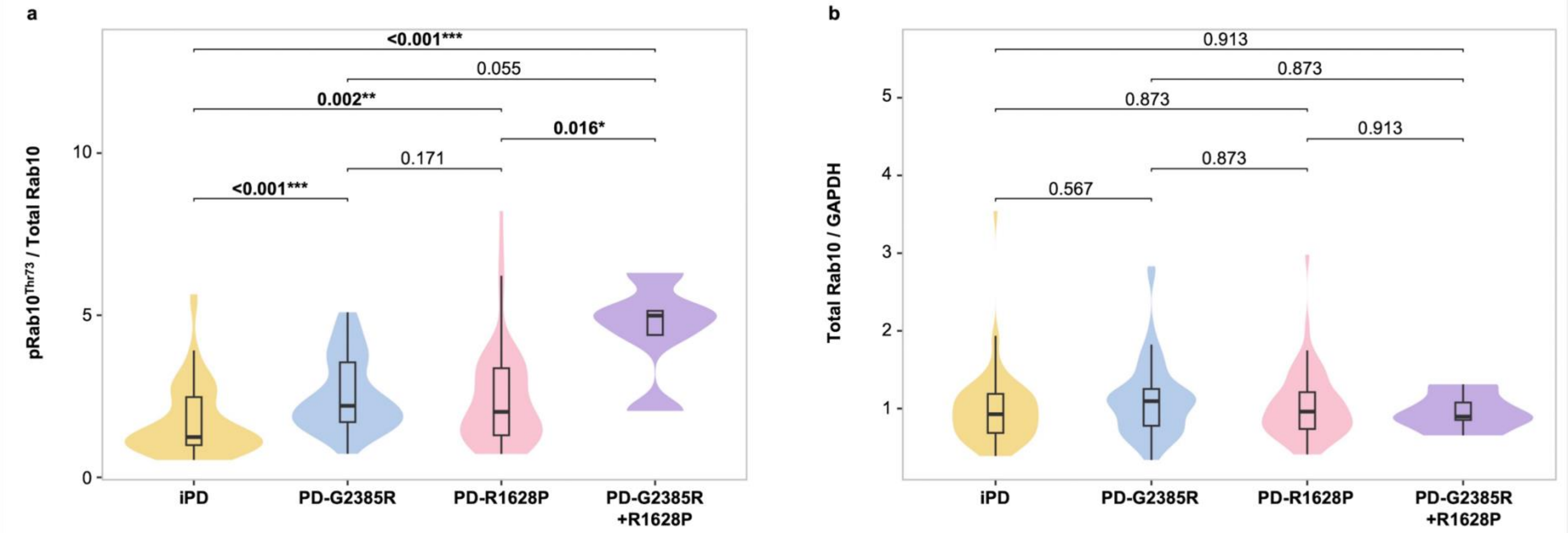


**a**

| | iPD | PD-G2385R | PD-R1628P | PD-G2385R+R1628P |
|---|---|---|---|---|
| **N** | 61 | 57 | 61 | 5 |
| **Median** | 1.24 | 2.21 | 2.02 | 4.99 |
| **IQR** | 1.48 | 1.84 | 2.07 | 0.75 |

**b**

| | iPD | PD-G2385R | PD-R1628P | PD-G2385R+R1628P |
|---|---|---|---|---|
| **N** | 61 | 57 | 61 | 5 |
| **Median** | 0.93 | 1.10 | 0.96 | 0.90 |
| **IQR** | 0.50 | 0.47 | 0.47 | 0.23 |

**Supplementary Figure 2: Comparisons of Rab10$^{Thr73}$ phosphorylation across PD subgroups stratified by *LRRK2* p.G2385R and p.R1628P status.**

Monocyte lysates were analyzed by quantitative immunoblotting. Quantified immunoblotting data are presented as ratios of (**a**) pRab10$^{Thr73}$/total Rab10 and (**b**) total Rab10/GAPDH, normalized to the average values obtained from an independent healthy control (experiment performed in duplicate). Pairwise comparisons between groups were conducted using the Kruskal-Wallis test with post hoc Dunn's test, with *p*-values adjusted using the Benjamini-Hochberg method (adjusted $p<0.05$). *$p<0.05$, **$p<0.01$, ***$p<0.001$. Abbreviations: iPD=patients with idiopathic Parkinson's disease; IQR=interquartile range; PD-G2385R=PD patients carrying the p.G2385R variant; PD-G2385R+R1628P=PD patients carrying both p.G2385R and p.R1628P; PD-R1628P=PD patients carrying p.R1628P.

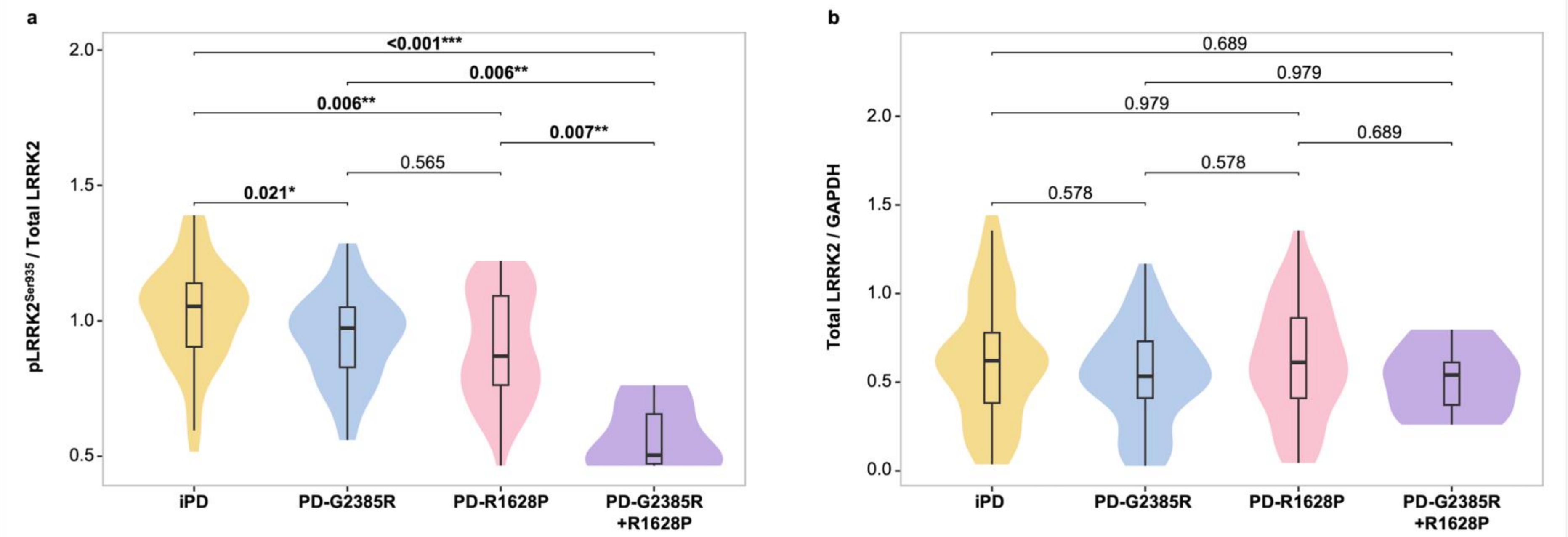


(a)

| | | | | |
|---|---|---|---|---|
| **N** | 57 | 54 | 54 | 5 |
| **Median** | 1.05 | 0.97 | 0.87 | 0.50 |
| **IQR** | 0.24 | 0.22 | 0.33 | 0.18 |

(b)

| | | | | |
|---|---|---|---|---|
| **N** | 57 | 54 | 54 | 5 |
| **Median** | 0.62 | 0.53 | 0.61 | 0.54 |
| **IQR** | 0.40 | 0.32 | 0.45 | 0.24 |

**Supplementary Figure 3: Comparisons of LRRK2$^{Ser935}$ phosphorylation across PD subgroups stratified by *LRRK2* p.G2385R and p.R1628P status.**

Monocyte lysates were analyzed by quantitative immunoblotting. Samples with faint or unquantifiable LRRK2 bands and total LRRK2/GAPDH values<0.02 were excluded from the analysis. Quantified immunoblotting data are presented as ratios of (**a**) pLRRK2$^{Ser935}$/total LRRK2 and (**b**) total LRRK2/GAPDH, normalized to the average values obtained from an independent healthy control (experiment performed in duplicate). Pairwise comparisons between groups were conducted using the Kruskal-Wallis test with post hoc Dunn's test, with *p*-values adjusted using the Benjamini-Hochberg method (adjusted $p<0.05$). *$p<0.05$, **$p<0.01$, ***$p<0.001$. Abbreviations: iPD=patients with idiopathic Parkinson's disease; IQR=interquartile range; PD-G2385R=PD patients carrying the p.G2385R variant; PD-G2385R+R1628P=PD patients carrying both p.G2385R and p.R1628P; PD-R1628P=PD patients carrying p.R1628P.

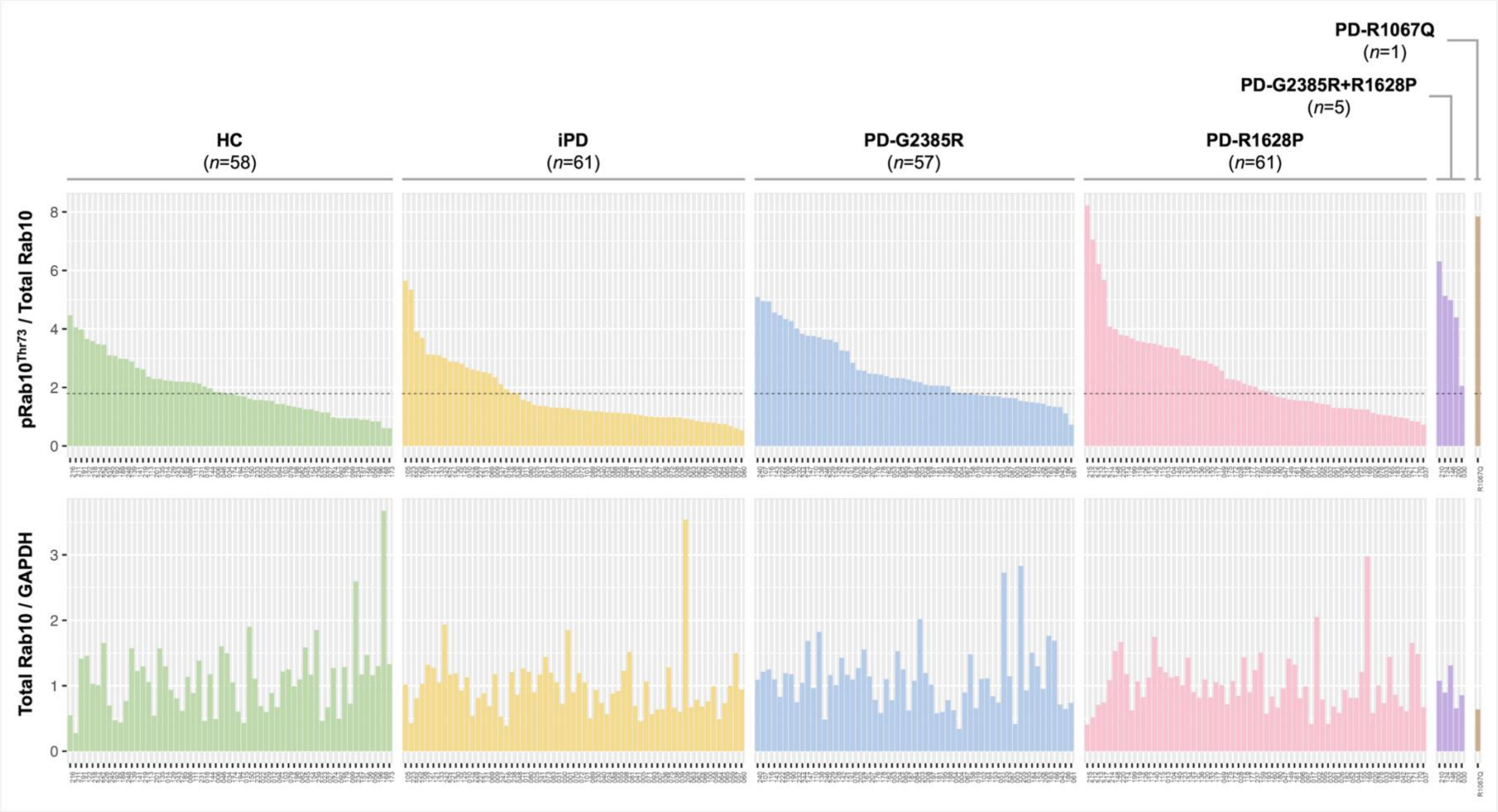


**Supplementary Figure 4: Distribution of pRab10$^{Thr73}$/total Rab10 and total Rab10/GAPDH levels for each study participant.**
The dotted horizontal line represents the median pRab10$^{Thr73}$/total Rab10 value in the HC group (median=1.79). Inter-individual variation was observed in pRab10$^{Thr73}$ phosphorylation, with all double-variant carriers (*n*=5/5), 68.4% of PD-G2385R (*n*=39/57), 54.1% of PD-R1628P (*n*=33/61), and 34.4% of iPD (*n*=21/61), showing levels above the median of HC. A manifesting carrier of the pathogenic *LRRK2* p.R1067Q variant was included as a positive control. Abbreviations: HC=healthy controls; iPD=patients with idiopathic Parkinson's disease; PD-G2385R=PD patients carrying the p.G2385R variant; PD-G2385R+R1628P=PD patients carrying both p.G2385R and p.R1628P; PD-R1067Q=PD patient carrying p.R1067Q; PD-R1628P=PD patients carrying p.R1628P.

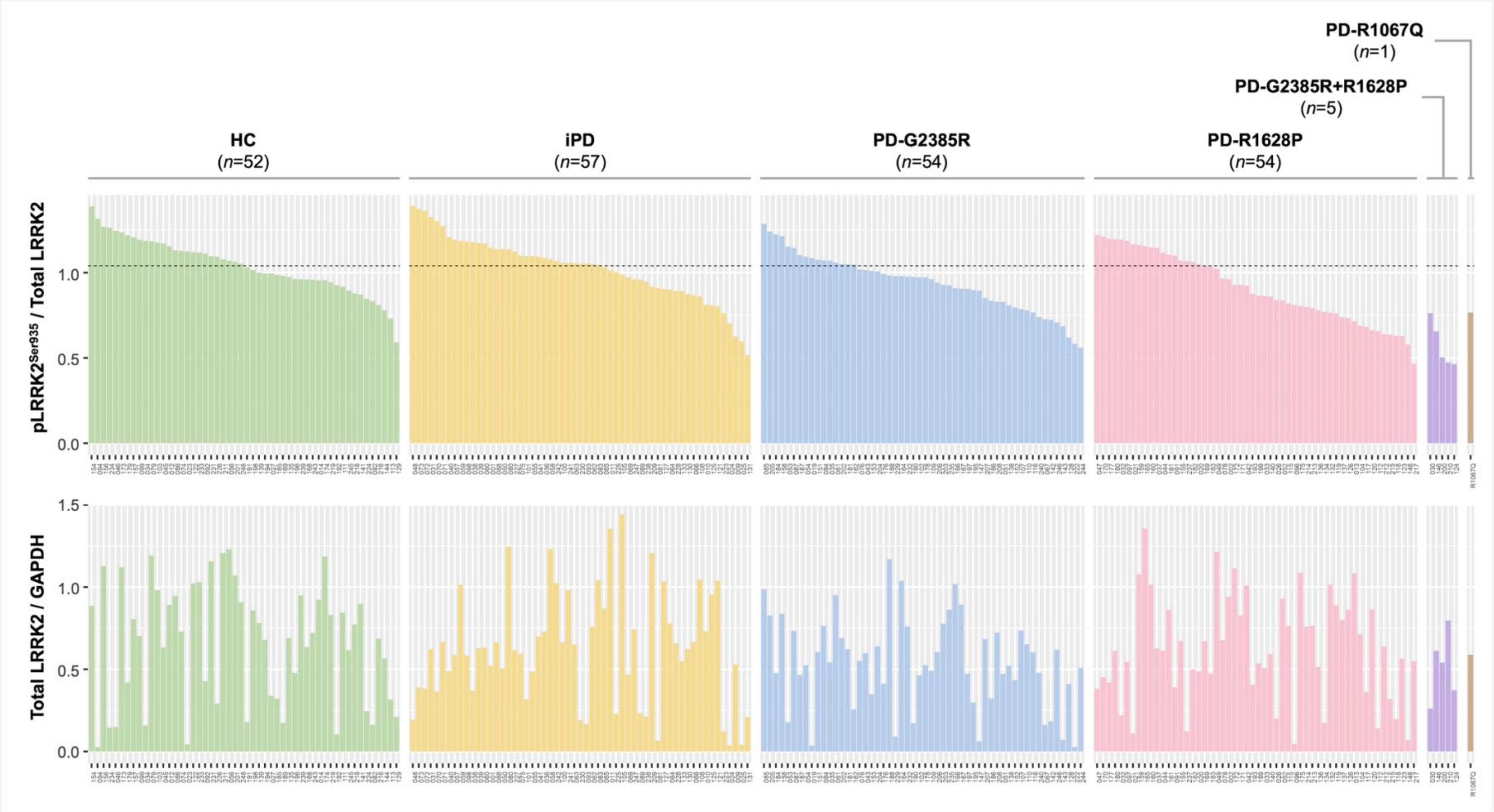


**Supplementary Figure 5: Distribution of pLRRK2$^{Ser935}$/total LRRK2 and total LRRK2/GAPDH levels for each study participant.** The dotted horizontal line represents the median pLRRK2$^{Ser935}$/total LRRK2 value in the HC group (median=1.04). A manifesting carrier of *LRRK2* p.R1067Q was included as a positive control. Inter-individual variation was observed in pLRRK2$^{Ser935}$ phosphorylation, with all double-variant carriers (*n*=5/5), 70.4% of PD-G2385R (*n*=38/54), 66.7% of PD-R1628P (*n*=36/54), and 43.9% of iPD (*n*=25/57) showing levels below than the median of HCs. Abbreviations: HC=healthy controls; iPD=patients with idiopathic Parkinson's disease; PD-G2385R=PD patients carrying the p.G2385R variant; PD-G2385R+R1628P=PD patients carrying both p.G2385R and p.R1628P; PD-R1067Q=PD patient carrying p.R1067Q; PD-R1628P=PD patients carrying p.R1628P.
